# Dynamics of Influenza Vaccination and Respiratory Virus Infections in Children: A Multistate Model Approach

**DOI:** 10.64898/2026.01.08.26343664

**Authors:** Anastasia Giorcelli, Vera Rigamonti, Marco Rocchi, Francesca Ieva, Vittorio Torri, Lara Cavinato, Shaun K Morris, Nicola Cotugno, Paolo Palma, Hedvig Nordeng, Nhung TH Trinh, Daniele Donà, Carlo Giaquinto, Costanza Di Chiara, Anna Cantarutti, Pedianet Network group

## Abstract

**Background:** Influenza vaccination prevents influenza and influenza-like illness (ILI) in children, but its potential influence on susceptibility to other respiratory viruses remains unclear. We aimd to evaluate the relationships between influenza/ILI, non-influenza respiratory virus (NIRV) infections, and influenza vaccination in children.

**Methods:** We conducted a retrospective cohort study using real-world data from the Italian Pedianet pediatric network, including children aged 6 months to 14 years followed during two influenza seasons (September 2022–April 2023 and September 2023–April 2024). Multistate models were used to describe transitions over time between vaccination and infection states (influenza/ILI and NIRV), allowing events to occur in any order and enabling estimation of transition probabilities and timing. A Self-Controlled Case Series (SCCS) analysis using Poisson regression estimated the incidence rate ratio (IRR) of NIRV infection associated with vaccination, accounting for influenza/ILI and seasonality.

**Results:** Among 91,695 children (median age 8 years; IQR 4–11; 51% male), vaccinated children showed delayed progression to NIRV infection and fewer early infections. Multistate models showed a substantially lower probability of transitioning to NIRV infection, by approximately 9- to >60-fold, in vaccinated vs unvaccinated children. In the SCCS analysis, influenza vaccination was associated with a reduced risk of NIRV infection (2022–2023:IRR 0.54; 95%CI,0.51–0.58; 2023–2024:IRR 0.60; 95%CI,0.56–0.64).

**Conclusions:** Influenza vaccination was associated with reduced NIRV infection risk in children, beyond its expected protection against influenza/ILI. These findings support virus interactions as a plausible mechanism shaping infection dynamics and reinforce the value of influenza vaccination in pediatric respiratory disease.

## INTRODUCTION

Influenza vaccination can effectively protect children from influenza/influenza-like illness (ILI) and its complications, significantly reducing global morbidity and mortality rates [1]. However, its effectiveness varies across seasons, depending on the antigenic match between the vaccine and circulating strains, often reducing the risk of influenza/ILI by 40-70% [2].

Although the influenza vaccine may protect against the risk of influenza/ILI, natural influenza may reduce the risk of non-influenza respiratory viruses (NIRV) infection by activating short-term non-specific immunity against other viruses [3]. However, the phenomenon of vaccine-associated virus interference remains poorly understood, leaving the potential impact of influenza vaccination on interactions with NIRV infections still unclear [4,5].

Virus interference, a concept recognized in virology for over six decades [6,7], refers to the phenomenon where prior infection with one respiratory virus modulates the infection and replication of another. This interplay is particularly relevant during concurrent respiratory virus outbreaks, as it may influence disease dynamics [8]. This interference can arise from direct interactions between viruses or from the initial virus modulating the host’s immune response, thereby affecting susceptibility to the second virus [6–8].

Some studies have suggested that influenza vaccination could increase the risk of NIRV infections, potentially due to the absence of the non-specific immunity typically induced by natural influenza [4,5,9,10]. On the other hand, other studies have found no link between influenza vaccination and an increased risk of NIRV infections [11,12]. However, the scientific literature on virus-virus-vaccine interference remains controversial due to design dependence, leading to a lack of evidence. Understanding the role of influenza vaccination in the context of influenza/ILI-NIRV interference is critical, as it may affect the seasonality and epidemiology of both influenza/ILI and other respiratory viruses, as well as influenza vaccination strategies during a future pandemic involving a non-influenza virus. To address this knowledge gap, this population-based cohort study aimed to assess the interference between influenza vaccination, influenza/ILI, and NIRV infections in children during two influenza seasons from September 2022 to April 2024.

## METHODS

### Setting

In Italy, community-based pediatricians provide free primary care for all children up to 14 years of age. This study used data from Pedianet, a well-established pediatric database that collects demographic, clinical, prescription, and vaccination information from more than 250 pediatricians, covering approximately 4% of the Italian pediatric population. Influenza vaccination is offered free of charge through participating pediatricians who record vaccination data in the Pedianet system. Pediatricians were classified as vaccination program participants based on national coverage thresholds. Further details on the Pedianet network, data structure, and vaccination program are provided in the **Supplementary Material eMethod**.

### Study design, aims, and population

We conducted a retrospective observational cohort study on children followed by pediatricians in the Pedianet network between September 2022 and April 2024. The study aimed to: (i) evaluate the transition times of children between different conditions – naïve (i.e., the health state at the beginning of the epidemiological season), experiencing influenza/ILI, having a NIRV infection, receiving influenza vaccination, or being disease-free – during each influenza season (September to April of the following year) (**Figure 1**), and (ii) assess the impact of influenza vaccination on the occurrence of NIRV infections across seasons. Eligible children were aged 6 months to 14 years, regularly followed by participating pediatricians, and under active care during the study period. A 15-day disease-free interval was defined following each acute episode. Further details on study design, definitions, and inclusion criteria are provided in the **Supplementary Material eMethod**.

**Figure 1.**
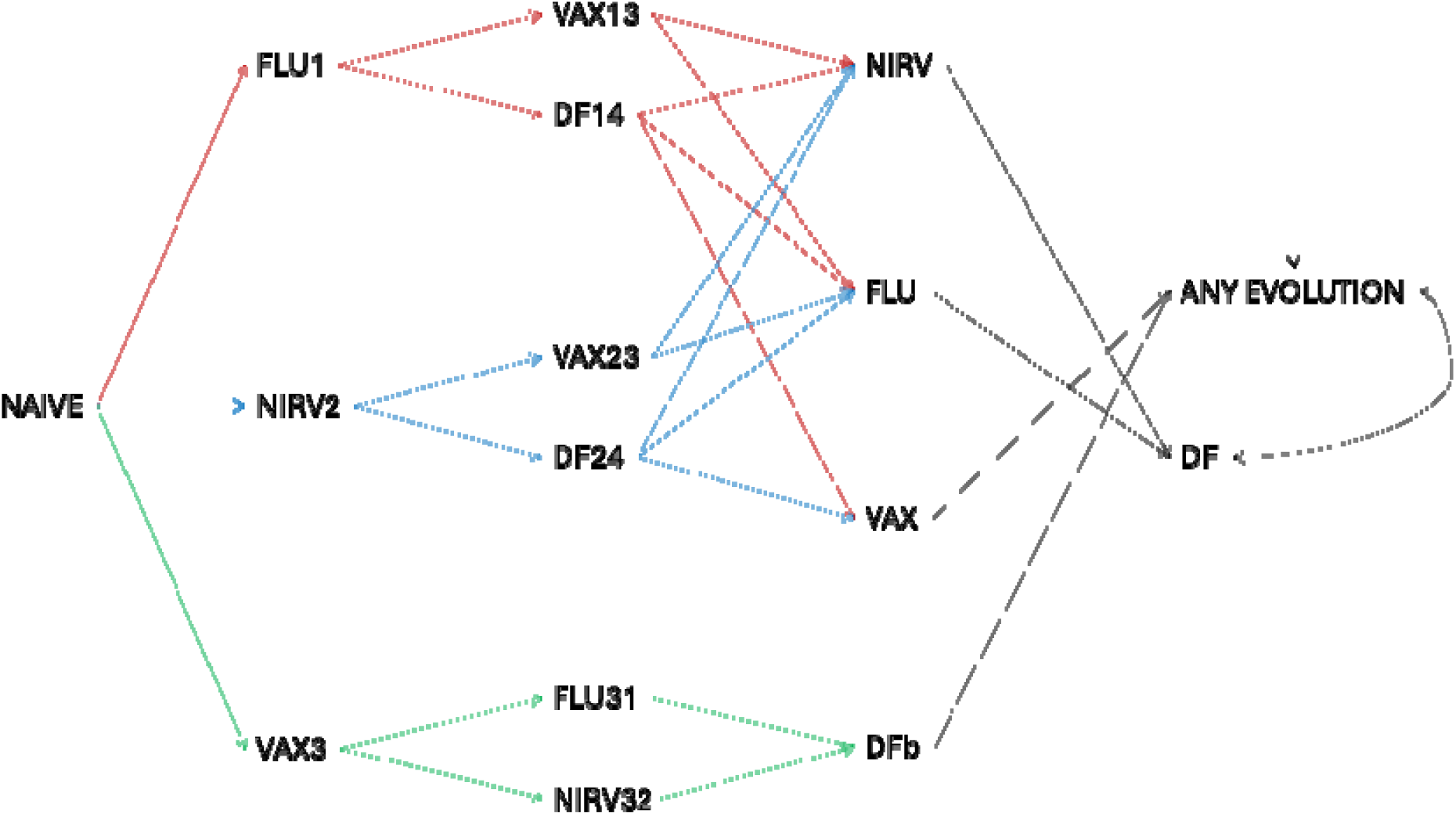
State transition diagram. Epidemiological season 2022-2024, Pedianet Network.

### Multistate model covariates

Covariates of interest included influenza vaccination, influenza/ILI, NIRV infection status, and corresponding time-to-event data. NIRV infections were identified using ICD-9-CM codes and free-text fields validated by a clinical data manager (**Supplementary Materials eTable 1**). Influenza vaccinations were identified using ATC codes (**Supplementary Materials eTable 1**), and children were considered vaccinated from 14 days after vaccination. An artificial intelligence algorithm based on NLP methods, previously described [13], was employed to identify influenza/ILI (**Supplementary Materials** **Appendix I**).

In cases where multiple influenza/ILI or NIRV infection records occurred within 15 days of the initial diagnosis, the subsequent records were considered follow-up visits. This classification was applied using a cleaning algorithm explained in a previous work [14].

### Measured confounders

A Directed Acyclic Graph (DAG) in DAGitty was used to define the interactions among the confounding factors included in the main analysis (**eFigure 2**). For each influenza season, the characteristic factors were: (i) children’s socio-demographic characteristics (i.e., age, sex, and area deprivation index [15]) and (ii) children’s health-related characteristics (i.e., medical exemptions). Exemption codes were used to retrieve information on chronic underlying conditions (**eTable 2**).

**Figure 2.**
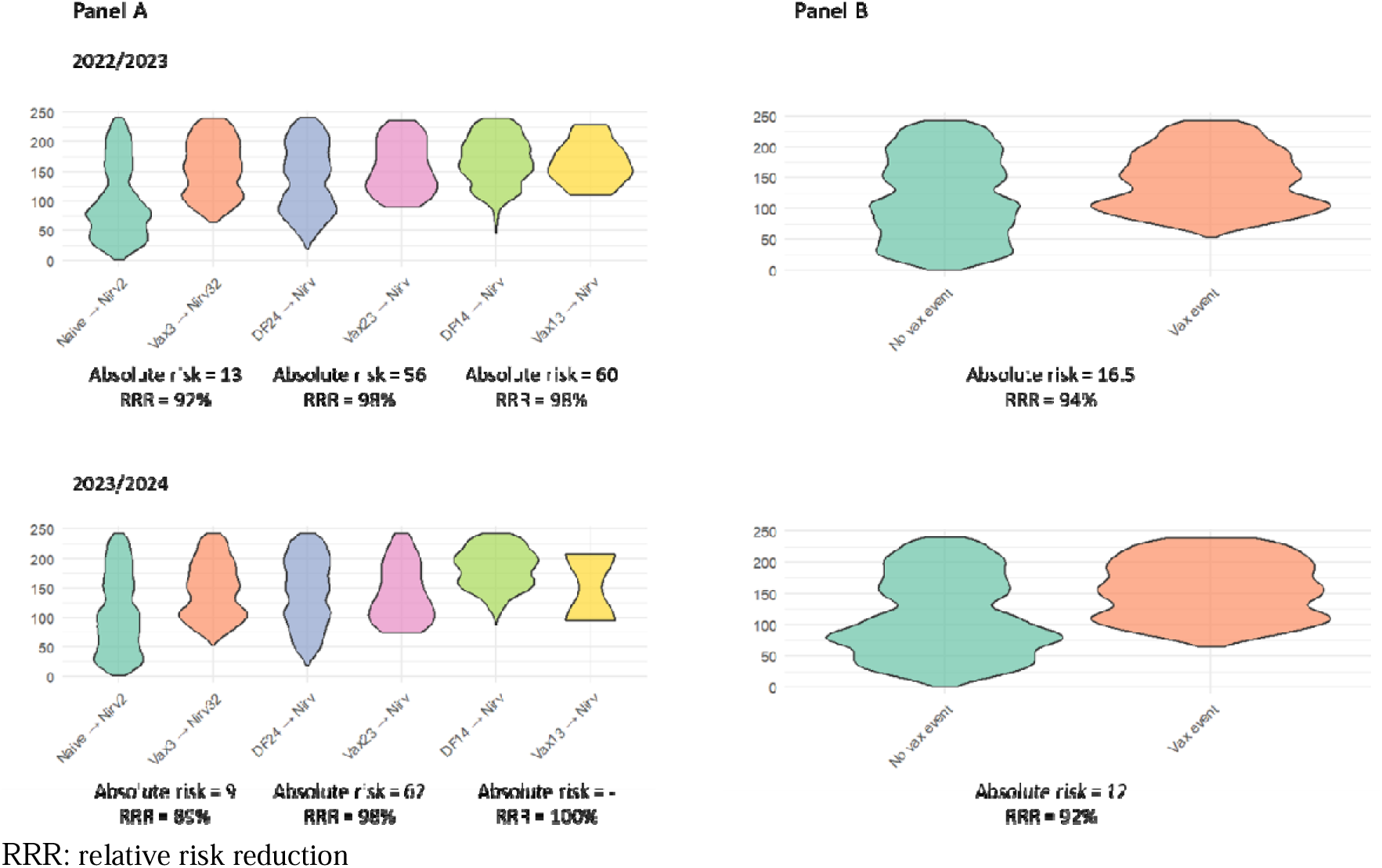
Timing and spread of transitions to NIRV infection across different pathways in the epidemiological seasons 2022/2023 and 2023/2024. Pedianet Network.

### Statistical Analysis

The multistate model was applied to explore longitudinal transition dynamics across influenza seasons, accounting for each child’s infection and vaccination history [16–18]. Within this framework, the imputed transition matrix enabled modelling of 16 possible transitions among five states: naïve, influenza-vaccinated, influenza/ILI-infected, NIRV-infected, and disease-free (**Figure 1**) [19]. Analyses were performed in R (packages mstate, dplyr, and ggplot2), using structured state mapping and transition extraction from the longitudinal dataset. For selected transitions, time-to-event distributions were computed and visualized with violin plots. The primary aim was to assess whether time to NIRV infection differed by prior influenza/ILI or NIRV infection and vaccination status. Specifically, we focused on transitions to NIRV infection, comparing children with and without prior vaccination, while accounting for previous infection history. Moreover, we assessed the probability, as well as the relative risk reduction, of transitioning to NIRV infection among vaccinated and unvaccinated children with the same history of prior transition(s).

We performed the analyses overall and by age groups (i.e., 6 mo. - 2 yr., 2 – 6 yr., 6 – 15 yr.). Further, a within-patient self-controlled case series (SCCS) design was adopted to assess the association between exposure to influenza vaccination and the onset of NIRV infection [20,21]. SCCS is a case-only approach aimed at eliminating between-persons confounding. In our application, children who experienced (1) both periods of exposure and non-exposure to influenza vaccination and (2) at least one NIRV infection were included. For each included patient, the observational time window began in September of each influenza season and ended at the end of that season (April). The observational time window was then divided into subperiods, with person-months of coverage and non-coverage for influenza vaccination. The exposure-outcome association was estimated using a conditional Poisson model to derive incidence rate ratios (IRRs) and their 95% confidence intervals (CIs), comparing NIRV infection rates during influenza vaccination coverage and non-coverage subperiods. Models were adjusted by age class, sex, deprivation index, and children’s underlying condition as time-fixed variables, and influenza/ILI as a time-dependent variable, with its effect remaining constant after the initial occurrence within a given influenza season. In addition, models accounted for interactions between vaccination and age, and between vaccination and exemptions. A fine calendar-time adjustment was modeled using natural cubic splines (6 df per season) to capture within-season variation in virus circulation **(Supplementary materials eFigure 3**). Sensitivity analyses evaluated the influence of prior infections and bacterial comorbidities (**Supplementary material eMethod**).

## RESULTS

A total of 91,695 children were included in the study, with 74,024 and 82,567 contributing to the 2022-2023 and 2023-2024 influenza seasons, respectively (**eFigure 1**).

In both the 2022-2023 and 2023-2024 seasons, most children were older than six years (65.43% in 2022-2023 and 65.77% in 2023-2024), with a median age of 8 years (interquartile range [IQR]: 4-11). A small proportion of children had underlying chronic conditions (4.64% in 2022-2023 and 4.52% in 2023-2024), while a high deprivation index was observed in 47.03% and 46.33% of children, respectively, during the 2022-2023 and 2023-2024 seasons (**eTable 3**).

### Time and risk of NIRV infection

Across both influenza seasons, vaccinated children consistently had a markedly lower probability of transitioning to NIRV infections than their unvaccinated peers. Depending on the pathway considered in the multistate model, this probability was reduced by approximately 9- to >60-fold, corresponding to a relative risk reduction (RRR) of ∼89–98%. Specifically, among children whose first state after naïve was vaccination, the probability of progressing to NIRV infection was about 9-to 13-fold lower (RRR ∼89–92%) than that of naïve, unvaccinated children across the two seasons. Among those whose first state after naïve was NIRV infection, children who subsequently transitioned into the vaccination state had a 56- to 62-fold lower probability of progressing to another NIRV infection (RRR ∼98%) compared with their unvaccinated counterparts. Finally, for children whose first state after naïve was influenza/ILI, those who transitioned into the vaccination state as their second state demonstrated an almost 89- to 100-fold lower probability of progressing to NIRV infections, corresponding to an RRR of ∼99% (**Figure 2, Panel A**).

**Figure 2** illustrates both the timing and spread of transitions to NIRV infection across different pathways in the epidemiological seasons considered. Children transitioning from a naïve state to NIRV infection (Naïve → NIRV2) showed earlier events with a widespread over time, whereas those whose first transition was vaccination (Vax3 → NIRV32) displayed later events with a narrower distribution. A similar pattern was observed for children transitioning from the disease-free state following a NIRV infection or influenza/ILI: transitions without vaccination (DF24 → NIRV and DF14 → NIRV) occurred earlier and were more widely distributed, while those with vaccination as the subsequent transition (Vax23 → NIRV and Vax13 → NIRV) were markedly delayed and more tightly distributed (**Figure 2** **– Panel A**). Overall, vaccinated children consistently demonstrated delayed and less frequent transitions to NIRV infection, with a narrower spread of timing compared with unvaccinated peers (**Figure 2** **– Panel B**). The stratified analysis by age group, along with sensitivity analyses, confirmed the main results (**Supplementary materials eFigure 3**). These results underline the effectiveness of influenza vaccination not only in delaying but also in reducing the risk of NIRV infection. Moreover, the findings highlight an important cross-interaction effect: children with a history of prior infections, whether influenza/ILI or NIRV, appeared to benefit even more from vaccination, showing an enhanced protective effect against subsequent NIRV infections. This suggests that prior exposure to respiratory pathogens may interact with vaccine-induced immunity, amplifying its protective role.

### Incidence Rate Ratio (IRR)

**Table 1** shows the risk of developing NIRV infection after influenza vaccination, taking into account the onset of influenza/ILI as a time-dependent variable, along with age, sex, deprivation index, children’s underlying conditions, and seasonal variation. The IRR for NIRV infection among vaccinated individuals compared to unvaccinated individuals was 0.54 (95% CI 0.51-0.58) in 2022-2023 and 0.60 (95% CI 0.56-0.64) in 2023-2024, indicating a significant reduction in risk of approximately 43%. Notably, our findings also demonstrated that the IRR for NIRV infection among those individuals who experienced influenza/ILI compared to naive individuals was 0.41 (95% CI 0.39, 0.43) in 2022-2023 and 0.38 (95% CI 0.36-0.41) in 2023-2024, indicating a lower risk of subsequent NIRV infection with a range of 59-62%. No significant association was observed between the risk of NIRV infections and age (**Table 1**). Sensitivity analyses restricting the cohort to (i) children without any NIRV infection in the previous influenza season and (ii) children with at least one NIRV infection in the previous influenza season were consistent with the main results (**Table 2**). Furthermore, the main results were also confirmed when only children without any bacterial NIRV infections were included during each influenza season (**Table 2**).

**Table 1.** The incidence rate ratio (IRR) and 95% confidence interval (CI) for NIRV infections among influenza-vaccinated children were estimated using Poisson regression within a self-controlled case series (SCCS) design. Epidemiological season 2022-2024, Pedianet Network.

| Condition | Seasons |  |
| --- | --- | --- |
|  | 2022-2023 | 2023-2024 |
|  | IRR [95% CI] | IRR [95% CI] |
| **Influenza vaccination** | 0.54 [0.51, 0.58] | 0.60 [0.56, 0.64] |
| **Influenza/ILI** | 0.41 [0.39, 0.43] | 0.38 [0.36, 0.41] |
| **Age class:** |  |  |
| **6m - <2y** | Reference | Reference |
| **2y - <6y** | 0.99 [0.95, 1.02] | 1.00 [0.96, 1.03] |
| **6y - <15y** | 0.99 [0.96, 1.02] | 0.97 [0.94, 1.01] |

m: months
y: years

**Table 2.** Sensitivity analyses of the incidence rate ratio (IRR) for NIRV infections among influenza-vaccinated children were conducted using Poisson regression within a Self-Controlled Case Series (SCCS) design. Epidemiological season 2022-2024, Pedianet Network.

| *No NIRV infection in the previous influenza season* |  |  |
| --- | --- | --- |
| **Condition** | **2022-2023 season** | **2023-2024 season** |
|  | **IRR (95% CI)** | **IRR (95% CI)** |
| Influenza vaccination | 0.51 [0.43, 0.60] | 0.56 [0.47, 0.66] |
| Influenza/ILI | 0.39 [0.35, 0.43] | 0.32 [0.28, 0.37] |
| Age class: |  |  |
| 6m - <2y | Reference | Reference |
| 2y - <6y | 1.00 [0.94, 1.07] | 1.00 [0.92, 1.08] |
| 6y - <15y | 1.01 [0.95, 1.06] | 0.99 [0.93, 1.06] |
| *At least a NIRV infection in the previous influenza season* |  |  |
| Influenza vaccination | 0.54 [0.50, 0.59] | 0.59 [0.55, 0.64] |
| Influenza/ILI | 0.41 [0.38, 0.44] | 0.39 [0.36, 0.42] |
| Age class: |  |  |
| 6m - <2y | Reference | Reference |
| 2y - <6y | 0.98 [0.94, 1.03] | 1.00 [0.96, 1.04] |
| 6y - <15y | 0.99 [0.94, 1.04] | 0.98 [0.94, 1.02] |
| *Any bacterial NIRV infection during each influenza season* |  |  |
| Influenza vaccination | 0.49 [0.44, 0.55] | 0.55 [0.51, 0.60] |
| Influenza/ILI | 0.34 [0.31, 0.37] | 0.33 [0.30, 0.36] |
| Age class: |  |  |
| 6m - <2y | Reference | Reference |
| 2y - <6y | 1.00 [0.95, 1.05] | 1.00 [0.95, 1.05] |
| 6y - <15y | 1.01 [0.96, 1.05] | 0.96 [0.92, 1.01] |

m: months
y: years

## DISCUSSION

This study represents the first application of multistate modelling to evaluate the interplay between influenza vaccination, influenza/ILI, and NIRV infections in a large pediatric cohort spanning two influenza seasons (2022–2024). By leveraging advanced longitudinal modelling techniques, our findings contribute to a deeper understanding of vaccine-virus interactions in children, providing novel insights into the broader benefits of influenza vaccination.

Our results demonstrated that vaccinated children had a markedly lower probability of transitioning to NIRV infection, with reductions of 9- to >60-fold corresponding to relative risk reductions of ∼89–99%. Transitions in vaccinated children also occurred later and with a narrower spread, whereas unvaccinated peers showed earlier and more dispersed events. Overall, vaccination was associated with both a substantial reduction and a delay in NIRV infections. These findings align with previous studies suggesting a protective association between influenza vaccination and NIRV infections [12], including those caused by respiratory syncytial virus (RSV), adenovirus, rhinovirus/enterovirus, and parainfluenza virus [22]. Notably, population-based studies have shown a reduction in RSV-related hospitalizations following influenza vaccination [23]. This could be potentially attributable to the development of nonspecific immunity induced by the influenza vaccine [24]. Conversely, some studies, albeit limited by small sample sizes, have suggested an increased risk of NIRV infections in vaccinated individuals compared to those recovering from natural influenza infection [4,9,10,25]. For instance, one study reported a four-fold increase in NIRV infections among vaccinated children [4]. However, the heterogeneity of findings across studies highlights the need for further large-scale investigations to reconcile these discrepancies. Our findings from the SCCS analysis further supported the protective role of influenza vaccination against not only influenza/ILI but also other respiratory viruses, identifying about a 56% reduction in NIRV infections among vaccinated children compared to their unvaccinated counterparts. This protective effect mirrors the observed 68-70% reduction in NIRV risk among children who have previously experienced influenza/ILI, underscoring the significance of virus-virus interactions and the potential for vaccines to mitigate viral burdens indirectly. Together, these findings emphasize the dual benefit of influenza vaccination: direct protection against influenza/ILI and indirect protection against other respiratory viruses [1,2]. With over 91,000 children included, geographically distributed across the nation, this study provides a robust foundation for assessing vaccine effectiveness and real-world interactions with viral infections. The extensive dataset ensures that the observed protective effects accurately reflect real-world dynamics, providing critical evidence for informed public health decision-making. Despite concerns over influenza vaccine mismatches observed in prior seasons, our findings demonstrate sustained vaccine effectiveness, including protection against NIRV infections, across both younger (0–<6 years) and older (6–<15 years) children.

A key strength of this study lies in its innovative design, which combines multistate modelling and SCCS approaches to account for the complexity of viral interactions and mitigate the impact of unmeasured confounders. The flexibility of the multistate model allowed for an accurate representation of real-world transition dynamics. At the same time, the large sample size ensured robust statistical power, even when analyzing high-dimensional transition matrices.

This analysis also presents some limitations. Diagnoses of influenza/ILI and NIRV infections were based on clinical data rather than microbiological confirmation, potentially leading to outcome misclassification. Importantly, outcome ascertainment differed between the two groups: influenza/ILI cases were identified using a validated algorithm, whereas NIRV infection cases were manually classified by the clinical data manager. This manual classification may have been more prone to misclassification, potentially resulting in under- or over-ascertainment of NIRV infection compared to influenza/ILI. In addition, reliance on clinical diagnoses, ICD coding, and NLP-based identification further introduces the possibility of misclassification, as these methods may lack sensitivity or specificity in identifying cases of interest. Taken together, these differences in ascertainment methods may have introduced bias; however, we believe this bias is unlikely to be relevant, as our findings are fully consistent with national surveillance data, making substantial misclassification improbable. Furthermore, the study did not include data on hospitalized patients or children followed by pediatricians who did not adhere to the influenza vaccination program, which may have influenced the magnitude of the observed effects. Residual confounding cannot be ruled out. Participation in Pedianet is voluntary, and only children of pediatricians adhering to the influenza vaccination program were included. This may reflect more health-conscious families and providers (healthy-vaccinee effect), possible confounding by indication, and differences in care-seeking behavior, although findings were consistent with national surveillance data. Moreover, the Pedianet dataset does not include information on non-pharmacological measures, such as hand hygiene and mask-wearing, which may have contributed to the observed reduction in NIRV infection risk following influenza vaccination. However, evidence from the COVID-19 pandemic suggests the possibility of risk compensation, with decreased adherence to prevention strategies among fully vaccinated individuals [26]. Additionally, the use of ICD-9-CM codes and free-text fields, even when validated by a clinical data manager and an NLP-based artificial intelligence algorithm, may be limited by variable coding practices, incomplete documentation, and center-specific terminology. These factors could reduce diagnostic specificity, leading to misclassification and affecting the algorithm’s reliability across different settings. Furthermore, in this study, we required at least one dose to consider a child full vaccinated. This definition may have introduced misclassification. In fact, two doses are recommended for children receiving influenza vaccination for the first time before nine years old. However, qualitative research, such as the study by Costantino et al. [27], suggests that caregivers vaccinating their children are likely to complete the initial two-dose regimen and, about our data, less than 10% of children were potentially misclassified as fully vaccinated, which could have biased our estimated towards the null. Lastly, while defining the season from September to April may limit the generalizability of results for respiratory viruses with less winter seasonality, such as enterovirus, the rapid decline in influenza vaccine-induced antibody titers within six months [28,29] suggests that the main interactions between influenza vaccination, influenza/ILI, and NIRV infections occur in the early post-vaccination period.

Given the novelty of our findings and their contrast with some previous studies [4], further investigation is warranted to explore the underlying biological mechanisms. The concept of temporary nonspecific immunity induced by influenza vaccination, particularly in protecting against NIRV infections such as RSV, requires validation through immunological studies. Additionally, large-scale observational cohort studies, randomized vaccine trials, and experimental human challenge studies could provide further insights into the dynamics of viral interference and the broader benefits of influenza vaccination.

## CONCLUSIONS

This study provides robust, real-world evidence of a protective effect of influenza vaccination on NIRV infections in children. By reducing the overall burden of respiratory infections, influenza vaccination plays a crucial role in preventing pediatric respiratory diseases, offering both direct and indirect benefits. These findings underscore the importance of annual influenza vaccination as a cornerstone of pediatric public health strategies.

## Funding

This work is supported by grants from the Italian Ministry of Education, University and Research (P20224MZE4), PNRR 2022-NAZ-0524 — PRIN 2022 under the National Recovery and Resilience Plan (PNRR), Mission 4, Component 2, Investment 1.1 – Call 1409/22: Covid-19 and Acute Respiratory Infections: the Clinical and Epidemiological Changes in the Pediatric Population (the CARICE project); CUP: H53D23007460001.

## Contributors Statement

Dr. Anastasia Giorcelli and Dr. Vera Rigamonti contributed to: conceptualization, methodology, formal analysis, data curation, investigation, visualization, and writing – original draft.

Dr. Marco Rocchi, and Profs. Francesca Ieva and Lara Cavinato contributed to: conceptualized and designed the multistate model, and Review & Editing.

Dr. Vittorio Torri contributed to: conceptualized and designed the artificial intelligence algorithms, and Review & Editing;

Profs. Shaun K Morris, Francesca Ieva, Nicola Cotugno, Paolo Palma, and Carlo Giaquinto contributed to: critically review of the work for important intellectual content, and writing – Review & Editing.

Dr. Hedvig Nordeng and Dr. Nhung Trinh contributed to: visualization, validation, and writing – Review & Editing.

Drs. Daniele Donà and Carlo Giaquinto contributed to: resources, and writing – Review & Editing. Dr. Costanza Di Chiara contributed to: conceptualization, investigation, validation, supervision, and writing – Review & Editing.

Dr. Anna Cantarutti contributed to: conceptualization, methodology, formal analysis, investigation, visualization, funding acquisition, supervision, writing – Review & Editing.

All authors reviewed, edited, and approved the final version of the manuscript, authorized its submission for publication, and agreed to be accountable for all aspects of the work.

## Data Availability Statement

The data used in this study cannot be made publicly available due to Italian data protection laws. The anonymized datasets generated and/or analyzed during the current study can be provided upon request from the corresponding author, upon written approval from the Internal Scientific Committee.

## Ethical statement

This study adhered to the principles of the Declaration of Helsinki. Participation in the Pedianet database required voluntary informed consent from parents or legal guardians. Data are encrypted and anonymized before use, in accordance with Italian law and the General Data Protection Regulation, and assigned a unique numerical identifier. To ensure data privacy, Pedianet researchers do not have access to the anonymization process and cannot identify individual patients. Approval of the study and access to the database was guaranteed by the Internal Scientific Committee of Società Servizi Telematici Srl, the legal owner of Pedianet.

## Acknowledgments

We wish to dedicate this work to the memory of Prof. Angela Lupatelli, whose valuable contributions and commitment to this study were deeply appreciated.

The authors gratefully acknowledge the head of Pedianet, Dr. Luigi Cantarutti, and the contributions of all the family pediatricians participating in Pedianet network: Fabiana Accardo, Arturo Alberti, Stefano Alboresi, Katya Alessio, Federica Alfani, Eva Alfieri, Sara Alfieri, Michela Alfiero Bordigato, Monica Aloe, Anna Aloisio, Angelo Alongi, Domenico Amabile, Flavia Amaro, Denise Amato, Elena Amodeo, Biagio Amoroso, Rosaria Ancarola, Barbara Andreola, Maria Luisa Andretta, Giampaolo Anese, Roberta Angelini, Maria Grazia Apostolo, Bruno Arcangeli, Giovanna Argo, Valentina Assirelli, Giovanni Avarello, Lucia Azzoni, Marta Bacciarini, Franco Balliana, Maria Carolina Barbazza, Maria Barberi Frandanisa, Patrizia Barbieri, Davide Bardella, Roberto Barone, Alice Bedini, Donatella Bellavere, Gabriele Belluzzi, Eleonora Benetti, Alessandra Beni, Chiara Maria Beretta, Fabio Berti, Roberto Bezzi, Filippo Biasci, Claudio Biondi, Giovanni Battista Biserni, Franca Boe, Ilaria Boiani, Stefano Bollettini, Francesco Bonaiuto, Emanuela Bonfigli, Anna Maria Bontempelli, Matteo Bonza, Gloria Borsari, Elisa Bortoli, Claudia Bortolin, Giuseppe Boscarelli, Lucia Boselli, Sara Bozzetto, Cecilia Bresci, Rosa Britta, Andrea Bruna, Ivana Brusaterra, Guido Brusoni, Mariella Bruzzese, Roberto Budassi, Massimo Caccini, Ilaria Cadel, Mariaclaudia Caiulo, Laura Calì, Maria Grazia Cammarata, Sonia Camposilvan, Laura Cantalupi, Luigia Caprio, Simone Carbogno, Chiara Cardarelli, Giovanna Carli, Sylvia Carnazza, Rita Casalboni, Anna Casani, Massimo Castaldo, Stefano Castelli, Serenella Castronuovo, Monica Cavedagni, Maria Silvia Cavinato, Cristina Cecamore, Stefania Censini, Giuseppe Egidio Cera, Chiara Chillemi, Rosa Maria Chiuri, Simona Ciccarelli, Francesca Cichello, Giuseppe Cicione, Niccolò Ciliani, Anna Giulia Cimatti, Anna Cingolani, Carla Ciscato, Mariangela Clerici Schoeller, Samuele Cocchiola, Eleonora Coclite, Margherita Codifava, Marta Cofini, Maurizio Coletta, Giuseppe Collacciani, Enrico Coltraro, Fabrizio Comaita, Ugo Alfredo Conte, Valeria Conte, Matteo Corchia, Francesca Corrias, Roberta Corro’, Rosaria Costagliola, Nicola Costanzo, Sandra Cozzani, Giulia Cremonini, Giancarlo Cuboni, Giorgia Curia, Valentino Curti, Salvatore Curto, Caterina D’alia, Vito Francesco D’Amanti, Michela D’Antoni, Antonio D’Avino, Alessandro D’Uva, Chiara Dalla Casa, Rita De Angelis, Roberto De Clara, Lorenzo De Giovanni, Annamaria De Marchi, Luisa De Marco, Chiara De Mutiis, Emanuele De Nicolò, Nicoletta De Polo, Irene Degrassi, Gian Piero Del Bono, Gigliola Del Ponte, Chiara Delehaye, Fabio Dell’Antonia, Giovanna Di Corcia, Tiziana Di Giampietro, Simona Di Loreto, Giuseppe Di Mauro, Francesco Di Mauro, Salvatore Di Palma, Anna Paola Di Renzo, Giuseppe Di Santo, Piero Di Saverio, Mattea Dieli, Marco Dolci, Mattia Doria, Stefano Drago, Dania El Mazloum, Giuseppe Elio, Maria Carmen Fadda, Clara Maria Faedi, Bernadette Faggioli, Pietro Falco, Elena Falcon, Mario Fama, Marco Faraci, Maria Immacolata Farina, Alessio Favali, Tania Favilli, Susanna Fedeli, Mariagrazia Federico, Michele Felice, Maurizio Ferraiuolo, Enrico Ferrara, Marta Ferrarese, Michele Ferretti, Mauro Gabriele Ferretti, Paolo Forcina, Patrizia Foti, Claudio Paolo Frattini, Luisa Freo, Ezio Frison, Fabrizio Fusco, Teresa Fusco, Alessandra Gabutti, Ambra Gagliardo, Giovanni Gallo, Roberto Gallo, Andrea Galvagno, Livia Garlisi, Stefano Gastaldo, Alberta Gentili, Pierfrancesco Gentilucci, Erica Giacomelli, Giuliana Giampaolo, Giuseppe Giancola, Francesco Gianfredi, Letizia Giaretta, Eugenia Giraldi, Silvia Girotto, Isabella Giuseppin, Laura Gnesi, Costantino Gobbi, Renza Granzon, Mauro Grelloni, Mirco Grugnetti, Silvia Gulden, Marwan Hamarneh, Martina Ielo, Giorgia Inchingolo, Giulia Innocenzi, Angela Cristina Intini, Antonina Isca, Urania Elisabetta Lagrasta, Maurizio Lanci, Massimo Landi, Maura Lazzari, Maria Rosaria Letta, Francesca Levi Della Vida, Giuseppe Lietti, Marianna Ligas, Cinzia Lista, Giuseppe Lorusso, Ricciardo Lucantonio, Elide Lucchi, Francesco Luise, Diego Luotti, Nadia Macropodio, Ivan Maddaluno, Matilde Maione, Tommaso Malusa, Elisabetta Manzali, Enrico Marano, Valeria Marchetti, Benedetta Marchi, Isabella Margherita, Francesca Marine, Lorenzo Mariniello, Pietro Marino, Gabriella Marostica, Valentina Marzetti, Sergio Masotti, Maura Mastrocola, Laura Mauri, Franco Mazzini, Clarissa Mazzotta, Rosa Maria Mele, Stefano Meneghetti, Annalisa Micheli, Francesca Miciotto, Massimo Milani, Stella Vittoria Milone, Antonella Minutoli, Donatella Moggia, Maria Chiara Molinari, Annalisa Monolo, Enrico Montagnani, Angela Maria Monteleone, Silvia Moretti, Giulia Maria Morresi, Angela Mortillaro, Annunziata Muggeri, Pierangela Mussinu, Carmen Muzzolini, Anna Naccari, Sara Nappini, Immacolata Naso, Novella Natale, Marina Navarra, Laura Nicoletti, Flavia Nicoloso, Erica Nistri, Monika Nitsch, Cristina Novarini, Laura Maria Olimpi, Riccardo Ongaro, Maria Maddalena Palma, Miriam Pambianchi, Angela Panariello, Vittorio Pandolfini, Stefano Pantano, Michael Panzenberger, Antonella Parlati, Enza Daniela Parrinello, Angela Pasinato, Andrea Passarella, Davide Pata, Viviana Dora Patianna, Pasquale Pazzola, Chiara Maria Pedrazzi, Monica Perin, Vanessa Perone, Cristina Perrera, Danilo Perri, Alberina Perrone, Sabrina Persia, Carla Alejandra Peruzzetto, Silvana Rosa Pescosolido, Giovanni Petrazzuoli, Giuseppe Petrotto, Vanna Piazza, Patrizia Picco, Elvira Pinelli, Ambrogina Pirola, Lorena Pisanello, Daniele Pittarello, Eleonora Polidoro, Roberto Ponti, Elena Porro, Adolfo Francesco Porto, Alfonsina Postiglione, Alfiero Prandoni, Elisabetta Profumo, Chiara Protano, Antonino Puma, Maria Paola Puocci, Anna Lucia Quitadamo, Ruth Raffeiner, Ferdinando Ragazzon, Giulia Reghelin, Paolo Regini, Andrea Righetti, Riccardo Righini, Rosaria Rizzari, Maria Oliva Rizzi, Enrica Romano, Cristiano Rosafio, Paolo Rosas, Rino Rosignoli, Matilde Rossi, Mariella Rossitto, Bruno Ruffato, Lucia Ruggieri, Francesca Rusalen, Annamaria Ruscitti, Annarita Russo, Pietro Salamone, Cristina Salvatori, Daniela Sambugaro, Francesco Emilio Sanfilippo, Luigi Saretta, Vittoria Sarno, Marcella Sasso, Renato Savastano, Valentina Savio, Antonio Scarcella, Ghislaine Sciarone, Nico Maria Sciolla, Antonio Scorrano, Maria Sellitto, Flavio Semenzato, Rossella Semenzato, Paolo Senesi, Martina Serafini, Daniela Maria Sgroi, Romina Silenzi, Carla Silvan, Giorgia Soldà, Martina Soliani, Cristina Spagnoli, Valter Spanevello, Sergio Maria Speciale, Sabrina Spedale, Francesco Speranza, Sara Stefani, Francesco Storelli, Gianni Tamassia, Paolo Tambaro, Anna Taveggia, Albino Terenghi, Luisa Toderini, Marco Todeschini Premuda, Giacomo Toffol, Gabriele Tonelli, Ilaria Tosetto, Miro Trebbi, Silvia Tulone, Angelo Giuseppe Tummarello, Antonella Ulliana, Cristina Vallongo, Sergio Venditti, Claudia Ventrici, Leonello Venturelli, Giulia Vigo, Maria Grazia Vitale, Sergio Vivarelli, Francescopaolo Volpe, Concetta Volpe, Barbara Vonella, Aldo Vozzi, Paola Wagner, Giulia Zanon, Chiara Zarbo, Maria Luisa Zuccolo.

## Conflict of Interest Disclosures (includes financial disclosures)

SKM have received funding from GSK, Pfizer, and Sanofi Pasteur for lectures and ad hoc advisory boards, none of which are related to this paper. The other authors have indicated that they have neither potential conflicts of interest nor financial relationships relevant to the article to disclose.

## Abbreviations

NIRV: non-influenza respiratory viruses
ILI: influenza-like illness
SPCS: Special Professional Commitment Services
FPs: Family Pediatricians
NLP: Natural Language Processing
SCCS: self-controlled case series
IQR: interquartile range
RRR: relative risk reduction
IRR: incidence rate ratio
95% CI: Confidence Interval at 95%
RSV: respiratory syncytial virus

## Notes

### Funding Statement

This work is supported by grants from the Italian Ministry of Education, University and Research (P20224MZE4), PNRR 2022NAZ0524 PRIN 2022 under the National Recovery and Resilience Plan (PNRR), Mission 4, Component 2, Investment 1.1 Call 1409/22: Covid-19 and Acute Respiratory Infections: the Clinical and Epidemiological Changes in the Pediatric Population (the CARICE project); CUP: H53D23007460001.

