## Supplementary materials for "Dynamics of Influenza Vaccination and Respiratory Virus Infections in Children: A Multistate Model Approach"

**eMethods.**

**Appendix I**

**eTable 1.** International Classification of Disease, Ninth Revision (ICD-9) and Anatomical Therapeutic Chemical (ATC). Definition of conditions.

**eTable 2.** Italian nationwide disease exemption codes.

**eTable 3.** Sociodemographic characteristics of the study population by influenza season.

**eFigure 1.** Eligible sample.

**eFigure 2.** Directed Acyclic Diagram representing the confounding variables acting in the model.

**eFigure 3.** Temporal Distribution of Infections and Influenza Vaccinations Across the Study Period.

**eFigure 4.** Timing and spread of transitions to NIRV infection across different pathways in the epidemiological seasons 2022/2023 and 2023/2024. Stratified analysis by age group, and among (i) children who experienced no NIRV infection in the previous influenza season, (ii) children who experienced at least one NIRV infection in the previous influenza season, and (iii) children with no history of bacterial NIRV infections.

**eMthods**

**Setting**

In Italy, community-based pediatricians within the National Health System provide free primary healthcare for children and adolescents aged 0 to 14 years.

The data for this study were retrieved from Pedianet (<http://www.pedianet.it>). This well-established pediatric database collects information on demographic characteristics, acute symptoms, inpatient and outpatient diagnoses (free-text or ICD-9-CM codes), drug prescriptions (Anatomical-Therapeutical-Chemical codes), specialistic visits, healthcare exemptions, and vaccinations. Data are provided by over 250 community-based pediatricians (referred to as pediatricians hereafter) who are members of the Pedianet network and use the Junior Bit® software in their clinical practice, caring for approximately 4% of the annual pediatric population in Italy. Data collected by pediatricians affiliated with Pedianet are updated monthly and stored in a centralized dataset in Padova, Italy [1].

The influenza vaccination program offers free vaccination through the voluntary participation of pediatricians, who provides influenza vaccination in their outpatient clinics. Vaccination data is recorded in the vaccination registry of the Pedianet database. Pediatricians participating in the program are reimbursed through the Special Professional Commitment Services (SPCS), which incentivizes accurate vaccination recording. For this study, pediatricians were classified as participants in the influenza vaccination program if they vaccinated at least 1.5% of their eligible patients (i.e., children aged six months or older). This threshold was established based on the average Italian seasonal influenza vaccination coverage rates [2]. Specifically, for children aged 6 months to 6 years, the coverage reaches approximately 14% and slightly decreases for those aged up to 14 years. This threshold also accounts for the average number of children followed by pediatricians, which is approximately 1,000 per practitioner.

**eStudy design**

Conditions were defined as follows: (1) naïve – status at the beginning of an epidemiological season for all children, defined as not yet vaccinated and free from influenza/ILI and/or NIRV infection; (2) influenza/ILI – children clinically diagnosed with influenza or ILI at a given point in time within each season; (3) NIRV infection – children clinically diagnosed with an acute non-influenza respiratory tract infection at a given point in time; (4) influenza vaccinated – children who received at least a single dose of the seasonal influenza vaccine at a given point in time; and (5) disease-free – a transitional state assigned to children following an influenza/ILI or NIRV infection. By design, overlapping diagnoses are not permitted; therefore, at 15 days after any infection diagnosis, the child automatically transitions into the disease-free status, indicating the resolution of the acute episode and the absence of new infections at that time (**Figure 1**).

Eligible children were included if they (i) were aged 6 months to 14 years, (ii) were followed by pediatricians who participated in influenza vaccination program [2], (iii) had regular routine child health visits [14], (iv) had at least one year of retrospective follow-up if older than one year, or six months if younger, and (v) were under care between September 1, 2022, and April 30, 2024. Details of the data selection for the two influenza season populations are provided in **eFigure1**.

**eSensitivity Analysis**

To understand if previous infections could impact vaccine-virus interactions, we conducted three sensitivity analyses. Specifically, for each influenza season, we repeated the time to NIRV infection and the SCCS design on (i) children who experienced no NIRV infection in the previous influenza season, (ii) children who experienced at least a NIRV infection during the previous influenza season, and (iii) children who had not experienced bacterial infections (i.e., pneumonia, sinusitis, pharyngitis, and otitis) across the influenza seasons to assess the potential influence of susceptibility to bacterial infections on risk of influenza/ILI and NIRV infections. Bacterial infections of interest were identified using ICD-9-CM codes and free-text fields validated by a clinical data manager, as specified above. In the subsamples (i) and (ii), we excluded the children younger than one year old as they did not have a complete follow-up in the previous influenza season.

[1] Cantarutti, A.; Gaquinto, C. Pedianet Database. In Databases for Pharmacoepidemiological Research; Springer: Cham, Switzerland, 2021; pp. 159–164.
[2] Di Chiara, C., Barbieri, E., Chen, Y. X., Visonà, E., Cavagnis, S., Sturniolo, G., Parca, A., Liberati, C., Cantarutti, L., Lupattelli, A., Le Prevost, M., Corrao, G., Giaquinto, C., Donà, D., & Cantarutti, A. (2023). Comparative study showed that children faced a 78% higher risk of new-onset conditions after they had COVID-19. *Acta paediatrica (Oslo, Norway : 1992)*, *112*(12), 2563–2571. <https://doi.org/10.1111/apa.16966>

**Appendix I.**

Influenza/ILI diagnoses were detected using an artificial intelligence algorithm that employed Natural Language Processing (NLP) techniques. This algorithm, previously described [15], captures all diagnoses recorded by pediatricians (i.e., free-text and ICD-9-CM codes: Influenza (487), influenza with pneumonia (487.0), influenza with other respiratory manifestations (487.1), influenza with other manifestations (487.8)) during routine clinical practice. Validation of the algorithm was conducted during the 2017-2018 influenza season in the Pedianet network, establishing a gold standard and demonstrating high performance. Subsequent confirmation processes were implemented across the 2022-2024 influenza seasons.

**eTable 1.** International Classification of Disease, Ninth Revision (ICD-9) and Anatomical Therapeutic Chemical (ATC). Definition of conditions.

| **Condition** | **ICD-9 codes** |
| --- | --- |
| Acute nasopharyngitis | 460 |
| Acute sinusitis | Acute sinusitis (461), acute maxillary sinusitis (461.0), acute frontal sinusitis (461.1), acute ethmoidal sinusitis (461.2), acute sphenoidal sinusitis (461.3), other acute sinusitis (461.8), acute sinusitis unspecified (461.9) |
| Acute pharyngitis | Acute pharyngitis (462) |
| Acute tonsillitis | Acute tonsillitis (463) |
| Acute laryngitis | Acute laryngitis (464.0), acute laryngitis w/o obstruction (464.00), acute laryngitis with obstruct (464.01), acute tracheitis no obstruction (464.10), acute tracheitis w obstruct (464.11), acute laryngotracheitis (464.2), acute laryngotracheitis no obstruction (464.20), acute laryngotracheitis with obstruction (464.21), acute epiglottitis (464.3), acute epiglottitis no obstruction (464.30), acute epiglottitis with obstruction (464.31), croup (464.4) |
| Acute otitis | Nonsuppurative otitis media and Eustachian tube disorders (381), Suppurative and unspecified otitis media (382), Mastoiditis and related conditions (383) |
| Acute bronchitis | Acute bronchitis (466.0) |
| Acute bronchiolitis | Acute bronchiolitis (466.1) |
| Pneumonia | Pneumonia (480-486) |
|  | **ATC codes** |
| Influenza vaccination | J07BB01, J07BB02, J07BB03, J07BB04, J07BB05 |

**eTable 2.** Italian nationwide disease exemption codes. Children without any exemptions have been categorized as healthy.

| **Disease** | **Disease Exemption Code** |
| --- | --- |
| Cystic fibrosis | 018 |
| Diabetes mellitus | 013 |
| Chronic obstructive pulmonary disease | 024, 057 |
| Asthma | 007 |
| Congenital and acquired immunodeficiency (including HIV) and/or immunosuppressive therapy | 003, 020, 048, 050, 052 |
| Neurological and neurocognitive conditions (including Down syndrome) | 011, 017, 038, 044, 065 |
| Prematurity | 040 |
| Renal failure | 023, 061 |
| Congenital cardiac disease (including heart failure) | 002, 021 |
| Chronic liver conditions | 008, 016 |

**eTable 3.** Sociodemographic characteristics of the study population by influenza season.

| **Characteristics** | | **Influenza season** | |
| --- | --- | --- | --- |
|  |  | **2022 - 2023**  **(N=74,024)** | **2023 - 2024**  **(N=82,567)** |
| **Age:** | 6m - <2y | 7,076 (9.56%) | 8,043 (9.74%) |
|  | 2y - <6y | 18,515 (25.01%) | 20,222 (24.49%) |
|  | 6y - <15y | 48,433 (65.43%) | 54,302 (65.77%) |
| **Age <1y** | Yes | 2,536 (3.43%) | 3,163 (3.83%) |
| **Sex:** | Female | 35,921 (48.53%) | 39,911(48.34%) |
| **Deprivation Index:** | High | 34,813 (47.03%) | 38,251 (46.33%) |
|  | Low | 25,892 (34.98%) | 27,570 (33.39%) |
|  | Missing* | 13,319 (17.99%) | 16,746 (20.28%) |
| **Underlying health related characteristics:**  **Cystic fibrosis**  **Diabetes mellitus**  **Chronic obstructive pulmonary disease**  **Asthma**  **HIV**  **Trisomy 21**  **Prematurity**  **Renal failure**  **Congenital heart disease**  **Chronic liver conditions** | Yes | 3,432 (4.64%)  11 (0.01%)  107 (0.14%)  11 (0.01%)  758 (1.02%)  138 (0.19%)  382 (0.52%)  2082 (2.81%)  15 (0.02%)  12 (0.02%)  4 (0.01%) | 3,732 (4.52%)  15 (0.02%)  123 (0.15%)  13 (0.02%)  705 (0.85%)  131 (0.16%)  370 (0.45%)  1428 (1.73)  15 (0.02%)  17 (0.02%)  4 (0.00%) |
| *Missing values are considered itself as a level because it includes both newborns without a defined deprivation index, or children with a mistaken/missing address  m: months  y: years | | | |

**eFigure 1.** Flow diagram illustrating the identification of the eligible population in the study.

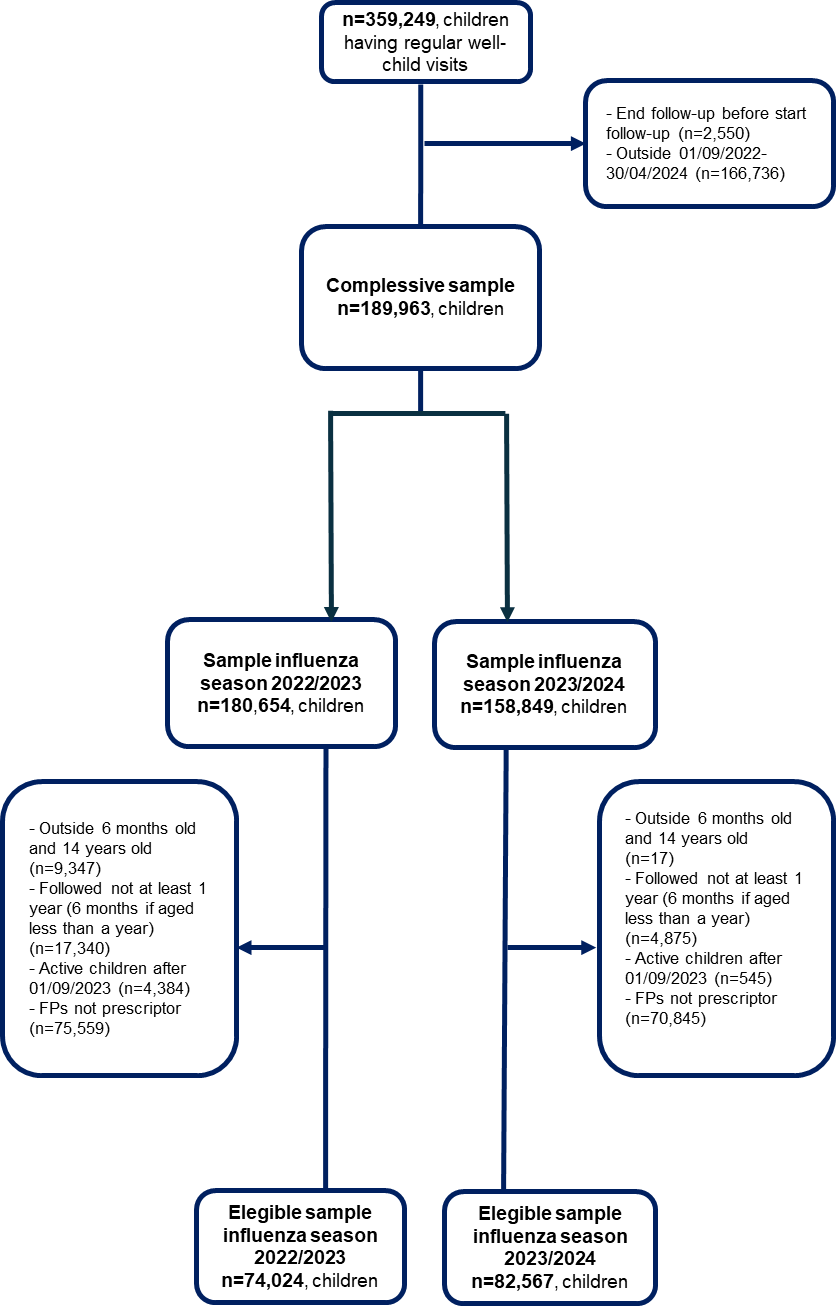

FPs: Family Pediatricians

**eFigure 2.** Directed Acyclic Diagram representing the confounding variables acting in the model.

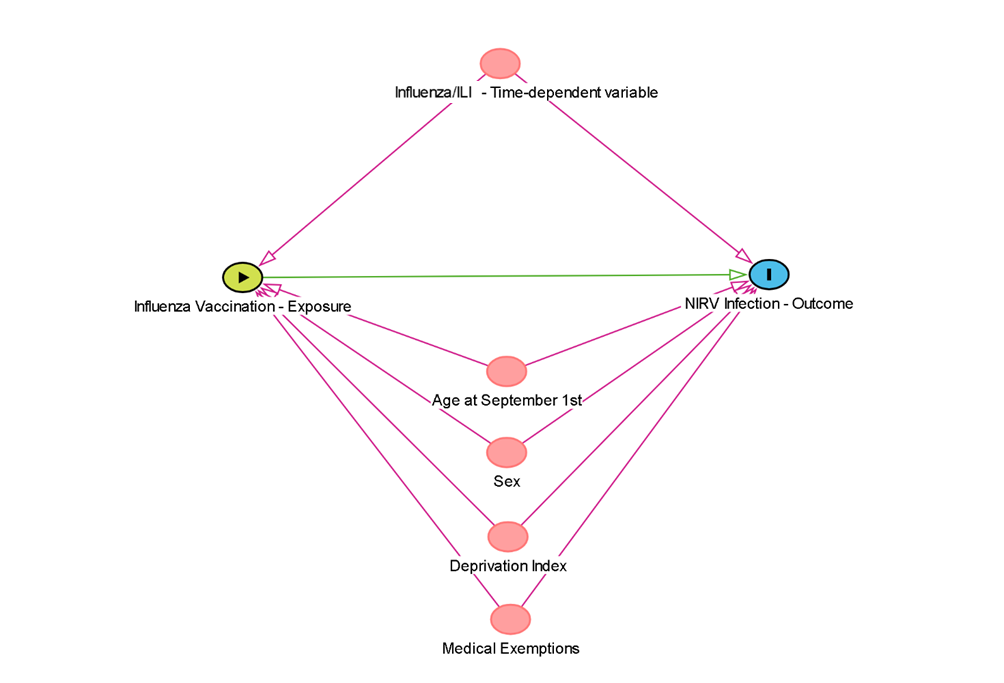

**eFigure 3.** Temporal Distribution of Infections and Influenza Vaccinations Across the Study Period.

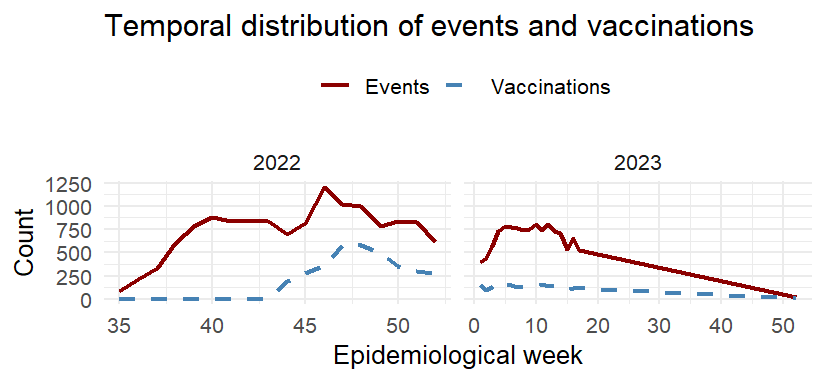

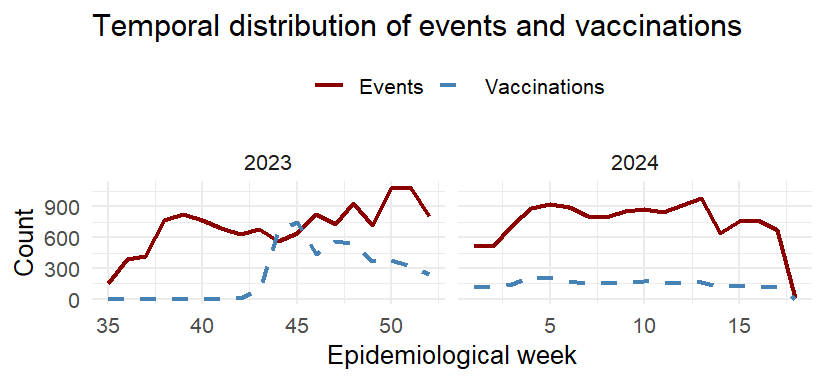

**eFigure 4.** Timing and spread of transitions to NIRV infection across different pathways in the epidemiological seasons 2022/2023 and 2023/2024. Stratified analysis by age group, and among (i) children who experienced no NIRV infection in the previous influenza season, (ii) children who experienced at least one NIRV infection in the previous influenza season, and (iii) children with no history of bacterial NIRV infections.

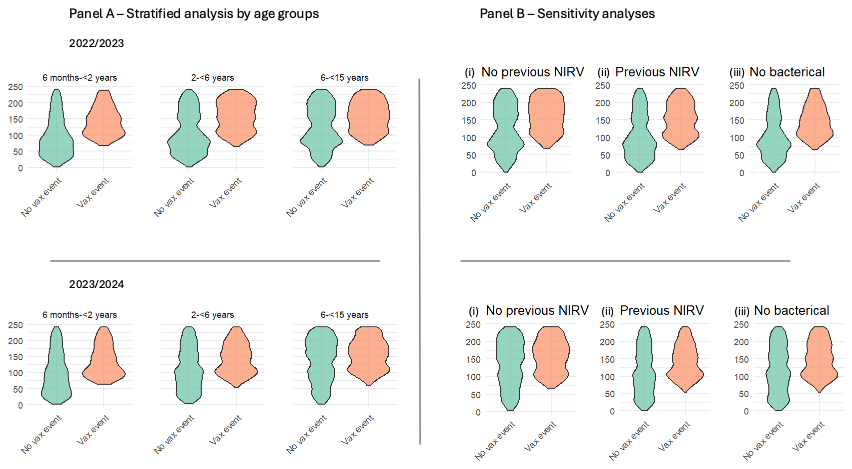

RRR: relative risk reduction
